# Analysis of COVID-19 case numbers: adjustment for diagnostic misclassification on the example of German case reporting data

**DOI:** 10.1101/2020.09.29.20203877

**Authors:** Felix Günther, Ursula Berger, Michael Höhle, Andreas Bender, Manfred Wildner, Iris M. Heid, Helmut Küchenhoff

## Abstract

**Background:** Reported COVID-19 case numbers are key to monitoring pandemic spread and decision-making on policy measures but require careful interpretation as they depend substantially on testing strategy. A high and targeted testing activity is essential for a successful Test-Trace-Isolate strategy. However, it also leads to increased numbers of false-positives and can foster a debate on the actual pandemic state, which can slow down action and acceptance of containment measures.

**Aim:** We evaluate the impact of misclassification in COVID-19 diagnostics on reported case numbers and estimated numbers of disease onsets (epidemic curve).

**Methods:** We developed a statistical adjustment of reported case numbers for erroneous diagnostic results that facilitates a misclassification-adjusted real-time estimation of the epidemic curve based on nowcasting. Under realistic misclassification scenarios, we provide adjusted case numbers for Germany and illustrate misclassification-adjusted nowcasting for Bavarian data.

**Results:** We quantify the impact of diagnostic misclassification on time-series of reported case numbers, highlighting the relevance of a specificity smaller than one when test activity changes over time. Adjusting for misclassification, we find that the increase of cases starting in July might have been smaller than indicated by raw case counts, but cannot be fully explained by increasing numbers of false-positives due to increased testing. The effect of misclassification becomes negligible when true incidence is high.

**Conclusions:** Adjusting case numbers for misclassification can improve this important measure on short-term dynamics of the pandemic and should be considered in data-based surveillance. Further limitations of case reporting data exist and have to be considered.

## 1 Introduction

In the acute COVID-19 pandemic, politics as well as public health and academic institutions worldwide are faced with the challenge of evaluating existing surveillance data like time series of reported case counts in real time. It is important to analyze and interpret this data taking into account all potential limitations and uncertainties, in order to maintain the highest possible confidence in generated insights. This is particularly important the longer the pandemic lasts and ongoing restrictions in public life also foster a growing wariness of people.

Interpreting daily reported COVID-19 case numbers is pivotal to gain insights into the state and dynamics of the current pandemic situation in different regions, but has several drawbacks. Problems can especially occur if the number of performed tests or the testing strategy change over time. The number of conducted PCR-tests has increased substantially in many European countries in summer 2020 [1], which coincided with an increase in observed case counts in July and August 2020, for example in Germany. In retrospect, this development of increasing case counts can be seen as a precursor of the strong second COVID-19 wave in Germany. At that time, however, it led in parts of society to increased skepticism whether the increasing case numbers were only false-positive test results and whether implemented measures to control the pandemic situation were superfluous.

There are three main challenges for the analysis and interpretation of reported case counts: first, the temporal assignment of the reported cases, second, misclassification in diagnostic tests, and third, a time-varying case detection ratio (dark figure). In this work we focus on the first two problems and refer to the third in the discussion.

To assess the short-term dynamics of an epidemic, it is common to look at the epidemic curve, defined as the number of disease onsets per day. Due to reporting delay, there are differences between date of disease onset and the date of case reporting and the time-series of newly reported cases can give a lagged and also in its structure incorrect impression of the acute pandemic situation. If data is collected on the day of disease onset, the epidemic curve can be constructed from this information. However, the reporting delay gives rise to occurred-but-not-yet-reported cases leading in real-time surveillance to a downward bias for days close to the current date. Utilizing individual-specific data on both disease onset and reporting date, it is possible to adjust for the reporting delay and to obtain an estimate of the epidemic curve based on nowcasting [2, 3, 4]. For SARS-CoV-2 and other pathogens, there exists the additional complexity that not all infected cases develop disease. Since the epidemic curve is a proxy for the number of exposed individuals over time (with a small time lag), it is still reasonable to assign those cases a synthetic disease onset date based on adequate assumptions.

Misclassification in COVID-19 diagnostics manifests in two different ways: infected persons who receive negative test results (false-negatives) and persons that are not infected but receive positive results (false-positives). One problem of false-negatives on the individual level is that infected persons are not aware of infection and not quarantined and can transmit the disease. On the population level, false-negatives lead to underestimating the number of infected individuals [5, 6]. False-positives lead on the individual level to superfluous quarantining and contact tracing, wasting time and resources. On the population level, false-positives lead to overestimating the number of infected individuals and could be the cause of intervention measures stricter than necessary [7]. In infectious disease surveillance, we are mainly interested in the population-level effects of misclassification. Since the impact of diagnostic misclassification depends on the number of tested persons and the true incidence, both changing over time, the apparent dynamics of reported case numbers can be misleading [8] and it is important to quantify the potential amount of distortion and adjust for it.

In this work, we provide an approach to adjust reported COVID-19 case counts for diagnostic mis-classification based on the matrix method [9]. We illustrate the impact of diagnostic misclassification on reported case counts on the example of Germany and the federal state of Bavaria under realistic assumptions for sensitivity and specificity of person-specific diagnostics and show that the approach can also be used to establish a lower-bound for the person-specific specificity. Furthermore, we illustrate how to use adjusted case numbers in downstream analyses like nowcasting or the estimation of the time-varying reproduction number. By this, it is possible to integrate solutions for both the problem of diagnostic misclassification as well as the problem of temporal assignment in the real-time analysis of case reporting data.

## 2 Methods

### 2.1 Data

Data on reported COVID-19 cases in Germany and Bavaria are collected based on the German Infection Protection Act (IfSG). For our analyses, we use daily German case numbers published by the Robert-Koch-Institute (RKI) [10], and, for Bavaria, the daily case numbers by the Bavarian Health and Food Safety Authority (LGL) [11]. In addition, the LGL provided us with person-specific case reporting data, including information on age, gender, and date of symptom onset if available (i.e., considered date of disease onset).

Furthermore, we use data on the number of SARS-CoV-2 laboratory tests that are directly reported by German and Bavarian testing facilities (university hospitals, research institutions, laboratories) to the respective health authorities. The testing facilities report the number of performed tests (analysed specimens) and the number of positive tests. The RKI publishes weekly data on the reported number of tests by German testing facilities [12, 13]. The LGL publishes daily numbers of laboratory tests and number of positive tests as reported by Bavarian testing facilities [11]. Because the data on case numbers come from a different source than the data on test numbers, the numbers of positive tests do not directly match to the number of reported cases and the reported number of performed tests does not directly correspond to the number of tested persons. Reasons for differences are diverse: (i) multiple testing of some individuals, (ii) reporting delay between the testing and the reporting of positive results from laboratories to local, regional and federal health authorities, and (iii) inconsistent reporting of test numbers by laboratories to the health authorities [14]. Furthermore, for the Bavarian data, persons whose tests are performed in Bavarian laboratories may not reside in Bavaria or vice versa.

Due to the weekly reporting of test numbers by the RKI, we focus our analysis on aggregated weekly case numbers for the German data. For Bavaria, we perform analyses based on daily data and make use of the person-specific information on disease onsets for estimating the epidemic curve.

In our main analysis, we focus on the time between May, 1 and Mid-September (utilizing data published on Sept 23, 2020, for Germany and data as per Sept 21, 2020, for Bavaria) as this was the most interesting phase of the pandemic in Germany with respect to changes in testing activity and their potential effects of misclassification on the epidemic curve, so far. Additionally, we provide current results for the analysis of the Bavarian data in the Supplemental Material and regular updates of the analysis on a public webpage (https://corona.stat.uni-muenchen.de/nowcast/).

### 2.2 Statistical approach to adjust reported case numbers for diagnostic misclassification

We present an approach to adjust reported case numbers in a given period (e.g., per day or week) for misclassification in the COVID-19 laboratory diagnostics (called examination in the following). Let *NT*_*t*_ be the number of examined persons whose test results would be reported to the health authorities on a given day (or within a given period) *t* in the event of a positive test. We denote the number of persons with a positive examination reported at time *t* as 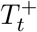 (observed cases). We now assume that the examination results might be error-prone and the actual number of cases at time *t* (of all examined persons during that period) is 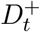 In practice, 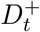 is unknown and we want to estimate it based on the observed case numbers 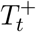, the number of examined persons *NT*_*t*_, and assumptions regarding the sensitivity and specificity of the person-specific examination. Based on elementary probability calculations, the (expected) number of observed cases can be expressed in terms of the true case numbers by

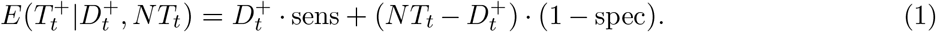

Equation (1) shows that the effects of a reduced sensitivity and specificity on the relation of observed case counts, 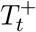, and true case counts, 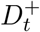, differ structurally: while a reduced sensitivity leads to an under-estimation of the true number of cases by the factor of the sensitivity, the effect of the specificity is additive and depends on the number of examined persons, *NT*_*t*_. To adjust the reported case numbers for misclassification we can re-order equation (1) and estimate the true number of cases at time *t* based on the reported case number, the number of examined persons, and the sensitivity and specificity:

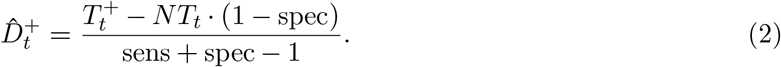

This estimator relates to the well known matrix method, see e.g., [9]. As described above, the number of examined persons, *NT*_*t*_, is not directly available in (German) case reporting data. We therefore approximate it using a statistical model based on the number of performed and the number of positive COVID-19 tests reported by laboratories and the number reported cases by the health authorities (cf. Supplemental Note 1).

### 2.3 Conceptualizing the sources of misclassification and deriving realistic person-specific misclassification probabilities

We conceptualize the sources of misclassification per PCR-test and per person-specific examination. For this, we searched for evidence on the extent of misclassification from literature and by consultation of experts from public health and virology. Besides the misclassification of PCR-tests under controlled laboratory conditions, we also consider misclassification from the following: (i) collecting and handling of specimens, (ii) additional aspects of uncertainty in PCR-tests under realistic conditions, e.g., due to timing of the test after infection or repeated testing in case of unclear results, (iii) varying testing strategy (physician-initiated/symptom-based vs. screening).

### 2.4 Adjusting reported case numbers for diagnostic misclassification

From the observed number of reported cases as published by the RKI (weekly numbers, Germany) or the LGL (daily numbers, Bavaria) and the estimated numbers of examined individuals, we estimate the true number of cases per time unit from equation (2) under various realistic misclassification scenarios. We compare results to reported case counts.

### 2.5 Estimation of the epidemic curve and the time-varying reproduction number based on adjusted case numbers

Daily reported case numbers are the basis for more complex downstream analyses, e.g., the real-time estimation of the epidemic curve (nowcasting) and estimation of the time-varying reproduction number, R(t). Those analyses provide a better characterization of the current state of the pandemic than reported case counts but require information on disease onset and reporting dates of cases [2]. To perform such analyses adjusted for diagnostic misclassification, we propose this general approach: (i) derive the adjusted number of reported cases per time unit, (ii) remove a randomly selected number of surplus reported (false-positive) cases per time unit from the data, and (iii) to estimate the epidemic curve and the reproduction number on the reduced data. Under the assumption of a low sensitivity, the number of misclassification-adjusted cases per time unit can be higher than the number of reported cases. We propose the following two-stage approach for the steps (ii) and (iii) from above and show the analytical validity (cf. Supplemental Note 1): (1) remove a given number of false-positive cases from the data based on the assumed specificity smaller than one and a sensitivity of one and estimate the epidemic curve based on these case counts and, (ii) adjust the resulting estimated epidemic curve for false-negatives based on an assumed sensitivity smaller than one. This procedure avoids the otherwise necessary up-sampling of data and cuts down considerably on computational resources.

We exemplify the misclassification-adjusted estimation of the epidemic curve and R(t) based on Bavarian case reporting data, since we were able to obtain person-specific information on disease onset for this German federal state.

### 2.6 Code and data

We provide R-code to reproduce our analyses on Github (https://github.com/FelixGuenther/mc_covid_cases_public) and update the results of the estimation of the misclassification-adjusted epidemic curve on our web page regularly (https://corona.stat.uni-muenchen.de/nowcast/). The proposed analysis can thus easily be extended to other countries or regions.

## 3 Results

### 3.1 Number of reported tests and reported cases in Germany and Bavaria

In the following, we refer to the time period between start of the laboratory reporting of conducted SARS-CoV-2 tests (calendar week 11 for Germany overall, March 16th for Bavaria) and end of our work’s observational period (as per calendar week 38 for Germany, Sept 20th for Bavaria), which is chosen to capture the low-level incidence in summer 2020. Based on the number of tests reported by laboratories, the testing activity in Germany started increasing in July and stabilized in September (Figure 1A) and this increase was more pronounced in Bavaria. Altogether, 15.7 million PCR-tests were reported by laboratories for Germany (i.e. 19 per 100 inhabitants) and 3.5 million by Bavarian laboratories (26 per 100). During spring/summer 2020, the number of reported positive tests (by laboratories) and reported cases (by health authorities) were the highest in the beginning of April, then decreasing to a low level in Mid-May and started rising in June/July (Figure 1B). The summer rise of case counts coincides with the increase of the test activity.

**Figure 1:**
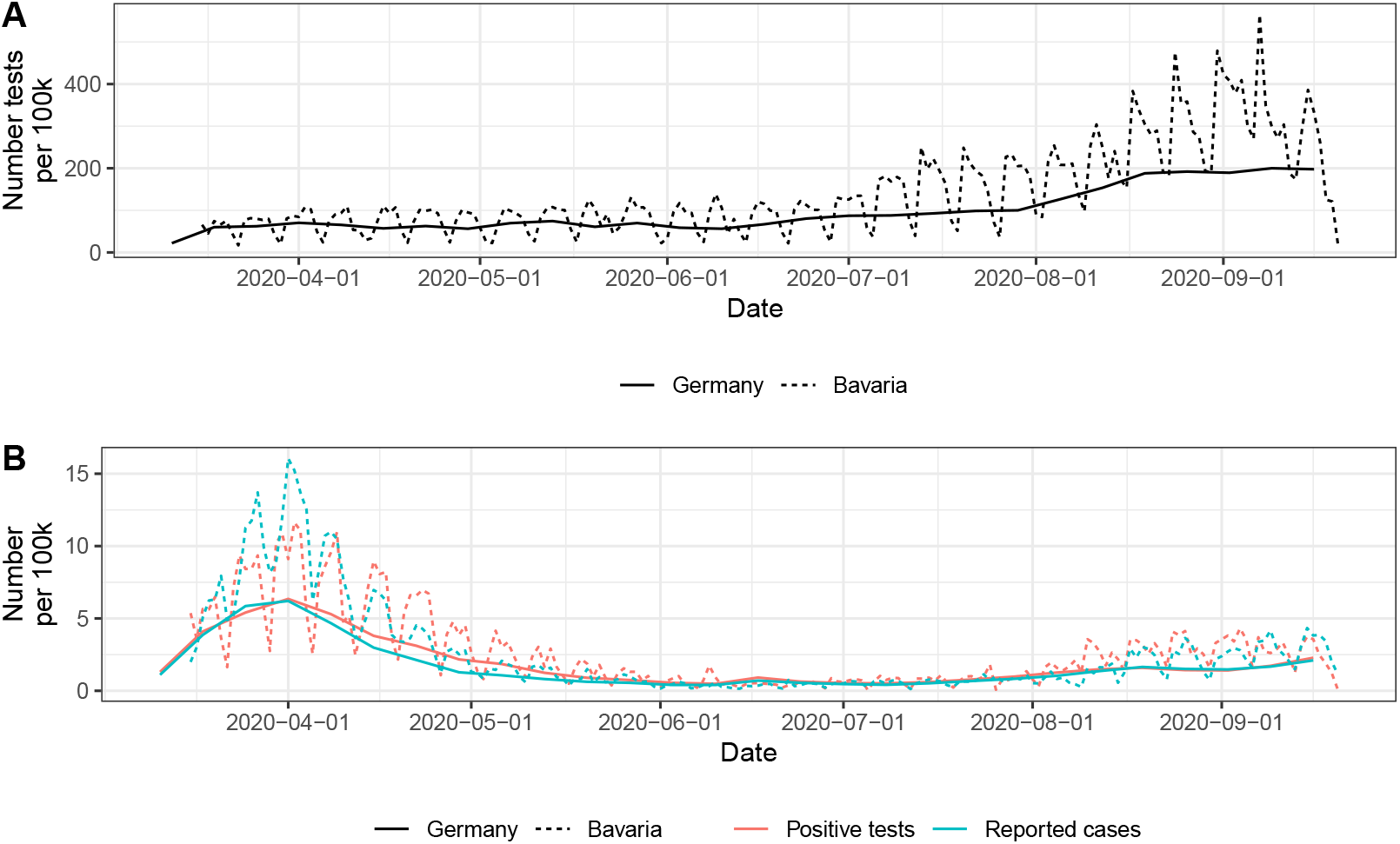
Reported number of performed PCR-tests and COVID-19 cases in Germany and Bavaria. Panel A shows the daily number of performed COVID-19 PCR-tests per 100,000 inhabitants in Germany and Bavaria as reported by the laboratories. Panel B shows the daily number of positive PCR-tests as reported by the laboratories and the number of COVID-19 cases as reported by the health authorities per 100,000 inhabitants. The daily numbers are average numbers from weekly reported data for Germany and daily reported numbers for Bavaria (with an obvious weekly cycle).

Note that the number of positive tests reported by laboratories and the number of COVID-19 cases reported by health authorities are not equal due to different reporting institutions. In Germany, there were 310,630 reported positive tests at the time of our analysis and 272,664 reported cases, in Bavaria 67,214 positive tests and 63,857 cases. Differences in time-series are particularly apparent in the daily data from Bavaria (Figure 1B): at the beginning of the first wave in February, more cases were reported by health authorities than positive tests by laboratories, most likely due to incompleteness of data reported from laboratories and incomplete coverage of reporting laboratories at this early phase. In our subsequent analyses, we focus on the time-period from May 1st to Sept 20th, 2020, where reporting of test numbers by laboratories was established and changes in the testing activity combined with low-level incidence in Germany is an ideal situation studying the impact of diagnostic misclassification.

### 3.2 Evidence on the performance of COVID-19 diagnostics

Routine COVID-19 examination in a screening or hospital setting is currently mostly done based on the detection of unique sequences of virus RNA using PCR-tests on clinical respiratory tract specimens of examined individuals [15]. Laboratories use different PCR-tests targeting different viral genes. The analytic sensitivity and specificity of PCR-tests applied to an adequately collected and handled specimen are generally reported to be very high [16]. In a proficiency test of German laboratories from April 2020, the authors found an average target-specific specificity between 97.8% and 98.6% and a sensitivity between 98.9% and 99.7% [17]. However, such numbers on the analytic performance of PCR-tests in laboratory settings do not directly relate to the person-specific performance of COVID-19 examinations in a hospital or screening setting, which is relevant for adjusting the reported case numbers in real-time surveillance. We systematically document sources of error in the person-specific COVID-19 examination (Table 1). While it is a substantial challenge to cover all aspects, some aspects should be noted in detail:

**Table 1:** Identified drivers of the performance of the person-specific COVID-19 examinations based on PCR-tests and their impact on sensitivity and specificity.

| Drivers of performance | Comment | Quantification, implication |
| --- | --- | --- |
| **Sensitivity** |  |  |
| Analytical | Very small error, depends on test | Average analytical sensitivity >98% |
| Specimens | Errors in collection and/or handling | Reduction due to damaged specimen, extent unclear |
| Real-world application | Test result depends on viral load | Reduction of sensitivity due to wrong timing of test: close to zero directly after infection, biggest ~8 days after infection (or shortly after/around symptom onset) |
| Testing strategy | Infection might be overlooked in screening due to wrong timing, individuals might get tested a second time after disease onset | Sensitivity higher in case of targeted testing or physician-initiated (e.g., due to symptoms): reduced sensitivity in screening application or testing of asymptomatic, sensitivity might change over time; infected individuals might get detected after repeated testing |
| **Specificity** |  |  |
| Analytical | Very small error, depends on test | Average analytical specificity >98% |
| Specimens | False-positives due to swaps or cross-contamination | Reduction due to cross-contamination, frequency unclear, rather low |
| Real-world application | Testing on dual targets and repeated testing in case of unclear results | Increase of specificity due to repeated testing and expert evaluation of results, extent might vary between laboratories |
| Testing strategy | Increased testing activity might reduce quality of testing due to limited resources | High workload in laboratories might decrease specificity of tests due to less time for validation of unclear results; might imply changes in specificity over time, extent of effect unclear |

With respect to the sensitivity, there are two factors that increase the probability of a false-negative examination result: first, inappropriate pre-analytical collection or handling of specimens, e.g., during transportation to laboratories, can lead to false-negative results. Second, the performance of PCR-tests in infected persons is reported to vary strongly depending on the time-point of the test after infection due to quantitatively insufficient viral RNA in the early pre-symptomatic phase [18]. The authors report a sensitivity close to zero directly after infection, an increase to 80% on day 8 (i.e., three days after typical symptom onset), and a decrease afterwards, all with high uncertainty. Similar results on the time-varying performance indicate a sensitivity of bigger 90% at the day of symptom onset with following decline [19]. Therefore, fast symptom-based testing should have a considerably higher sensitivity than testing in a screening setting. Multiple tests at different time-points (e.g., after symptom onset for a case with initial negative finding in a pre-symptomatic screening) can increase the person-specific sensitivity. It is difficult to quantify the overall sensitivity of the person-specific COVID-19 examination in the general testing regime, but it is certainly much lower than the high analytic sensitivity reported for tests in a controlled laboratory experiment. We perform our analyses based on an assumed sensitivity of 70% and 90% as a range of realistic values that also correspond to previously reported findings [20, 21, 22].

With respect to the specificity of the person-specific examination, chance or cross-contamination between specimens or swaps between infected and not infected individuals can lead to false-positive test results. It is recommended that tests are repeated to ascertain unclear results and to use tests that target two viral genes, which decreases the probability of false-positives considerably compared to a single PCR-test and/or single target tests [23]. However, the specific approach might depend on the laboratory and the overall situation (e.g., workload). Based on the Bavarian data, we show that a specificity for the person-specific examination lower than 99.5% is not empirically supported: a lower specificity would imply more false-positive cases than cases reported in the low-incidence phase in June and beginning of July, given the estimated number of examined persons in the respective time period (cf. section Adjusted case counts). For our analyses, we thus apply a specificity of 99.9%, 99.7% or 99.5%.

### 3.3 Number of examined persons in Germany and Bavaria

To adjust reported case counts for misclassification, we first derive the number of examined individuals per time unit (weekly or daily) based on the best performing varying-coefficient regression model to relate the number of reported cases to the number of reported positive PCR-tests over time (Supplemental Note 2, Supplemental Figure 1).

We find that, for the German data, the number of examined persons increases over time, most notably in the first half of August, and reaches a plateau in September (Figure 2A). The number of reported PCR-tests (derived from weekly data) matches the estimated number of examined persons quite closely. For the daily Bavarian data, we find a similar increase in examined persons, but larger differences in the number of reported PCR-tests by Bavarian laboratories and examined persons residing in Bavaria (Figure 2B). This difference is most pronounced at the end of German summer holidays (end of July to early September, where many inhabitants from other federal states were tested for free in Bavarian testing facilities upon travelling back home from the South through Bavaria). In the last days of our observation period here, reported numbers of PCR-tests decrease, which reflects the reporting delays between the time of the tests and the reporting of tests to health authorities. This reporting delay is also present in the case reporting data with respect to the number of reported cases per day by the local health authorities and we adjust for the reporting delay based on the nowcasting described below (cf. section Estimation of the adjusted epidemic curve).

**Figure 2:**
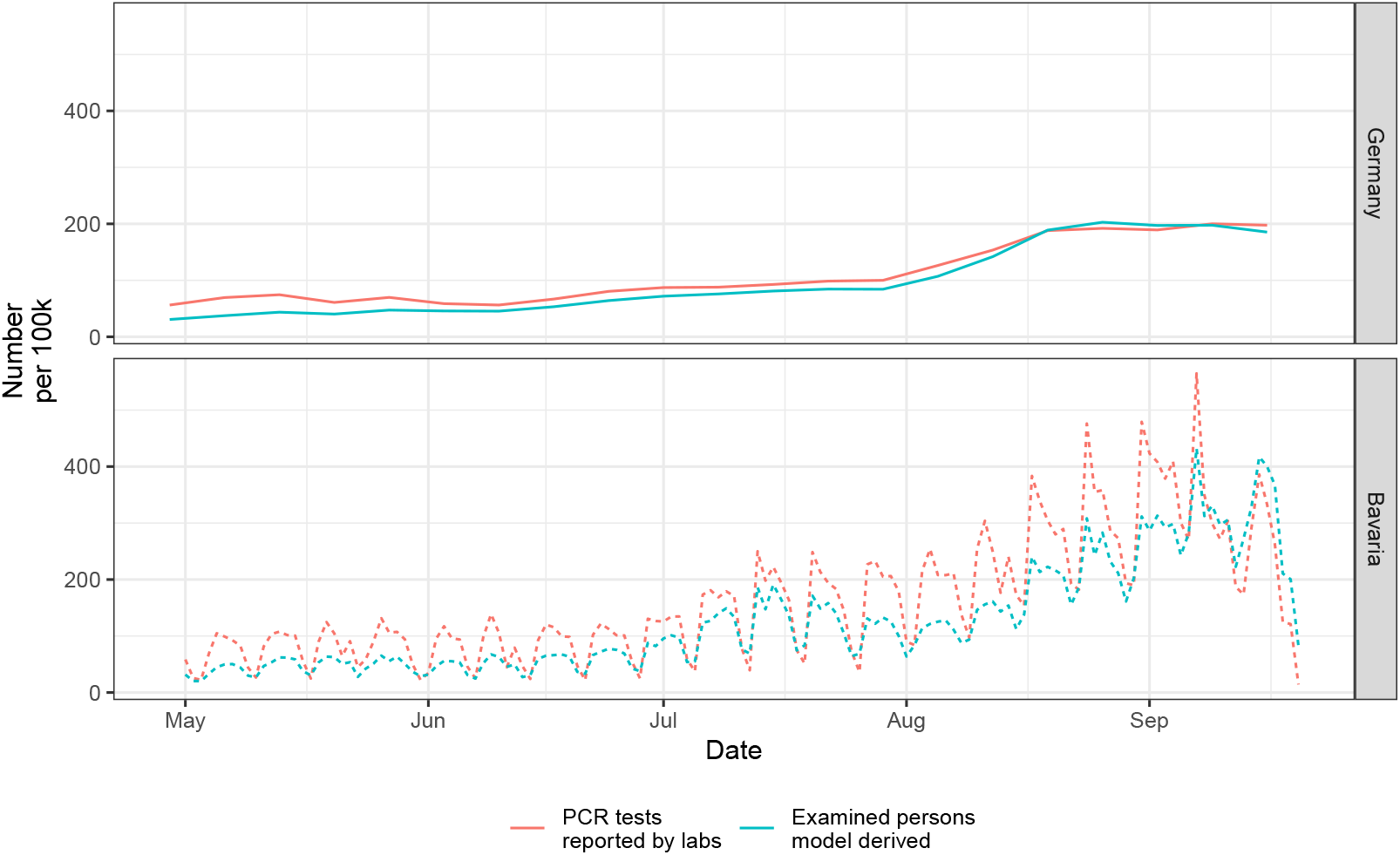
Reported number of PCR-tests and model-based number of examined persons in Germany and Bavaria. Shown are the daily numbers of performed PCR-tests per 100,000 inhabitants as well as the estimated number of examined persons. The estimated numbers of examined persons result from a varying-coefficient model fitted based on the association of the number of reported positive PCR-tests and the number of reported COVID-19 cases. Results for Germany stem from modeling weekly data and dividing by seven to illustrate results on a daily scale; the results for Bavaria are based on daily data (with an obvious weekly cycle).

### 3.4 Misclassification-adjusted case counts

Based on the derived numbers of examined persons in Germany and Bavaria, we adjust observed case numbers for misclassification in the person-specific diagnostic examination for SARS-CoV-2 infection under the assumptions of a sensitivity of 90% or 70% and a specificity of 99.9%, 99.7%, or 99.5% (Figure 3). Adjusting for a specificity less than one leads to a reduction of case numbers and adjusting for a sensitivity less than one increases the case numbers, as expected from theory. The impact of sensitivity and specificity on adjusted case counts is structurally different: while adjusting for imperfect sensitivity corresponds roughly to a time-constant upscaling of case numbers by the factor 1/sensitivity, the impact of the specificity varies over time due to varying numbers of examined individuals. The relative effect of adjusting for false-positives is the largest during July, when reported case numbers were low and testing activity started rising (Figure 4). The relative effect diminishes in August/September despite the increased testing activity during that time period due to an increased incidence.

**Figure 3:**
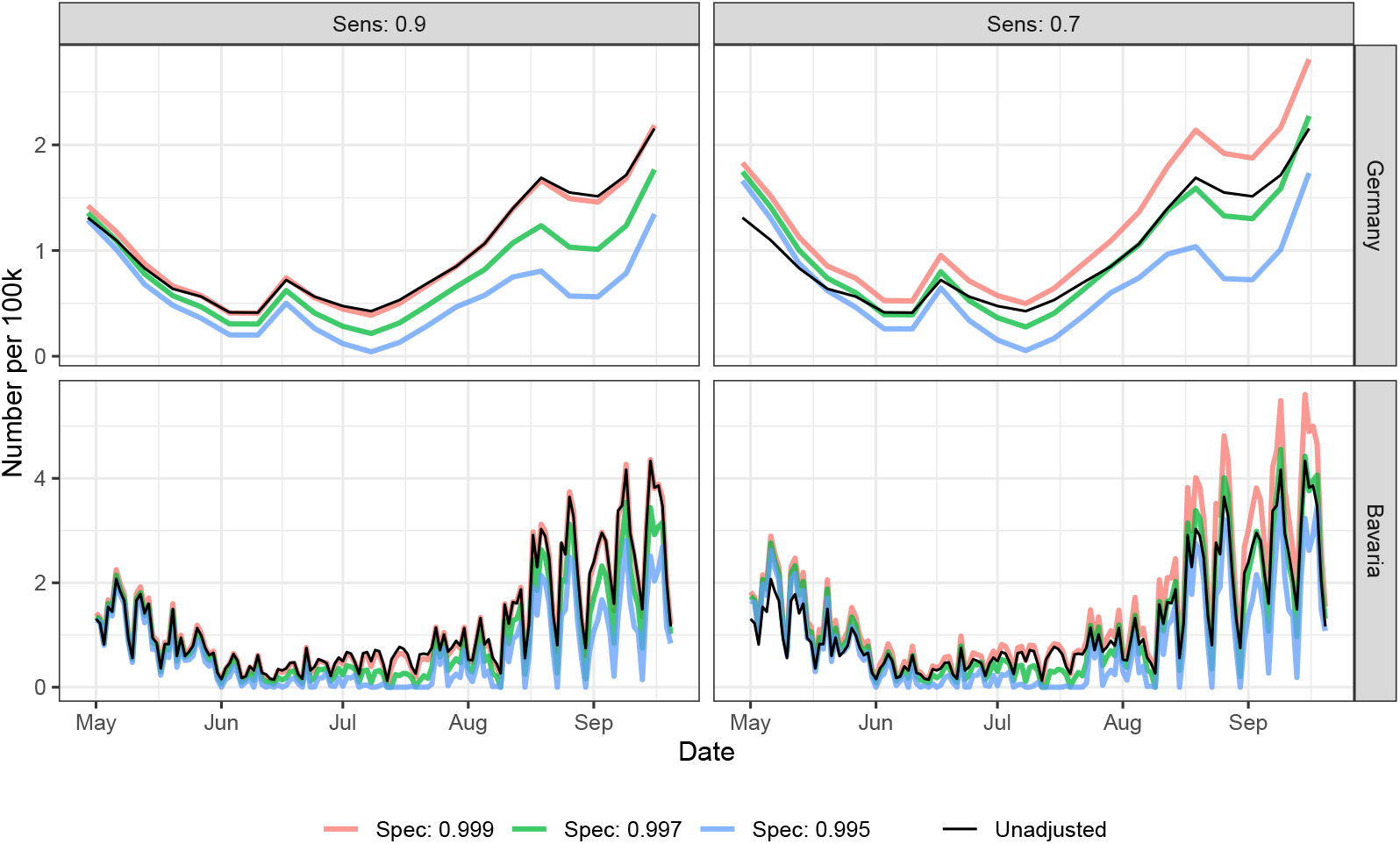
Misclassification adjusted case numbers for Germany and Bavaria. Shown are daily misclassification adjusted case numbers per 100,000 inhabitants for different assumptions regarding the sensitivity and specificity of the person-specific COVID-19 examination and the unadjusted reported case numbers. The smaller the assumed specificity, the higher the number of false-positives in the reported cases and the bigger the reduction of adjusted case numbers. The smaller the sensitivity, the bigger the probability of false-negative examinations and the higher the adjusted case numbers. Results for Germany stem from weekly reported data, the results for Bavaria are based on daily data (with an obvious weekly cycle). Note the different scales on the y-axis for German and Bavarian data for better visibility.

**Figure 4:**
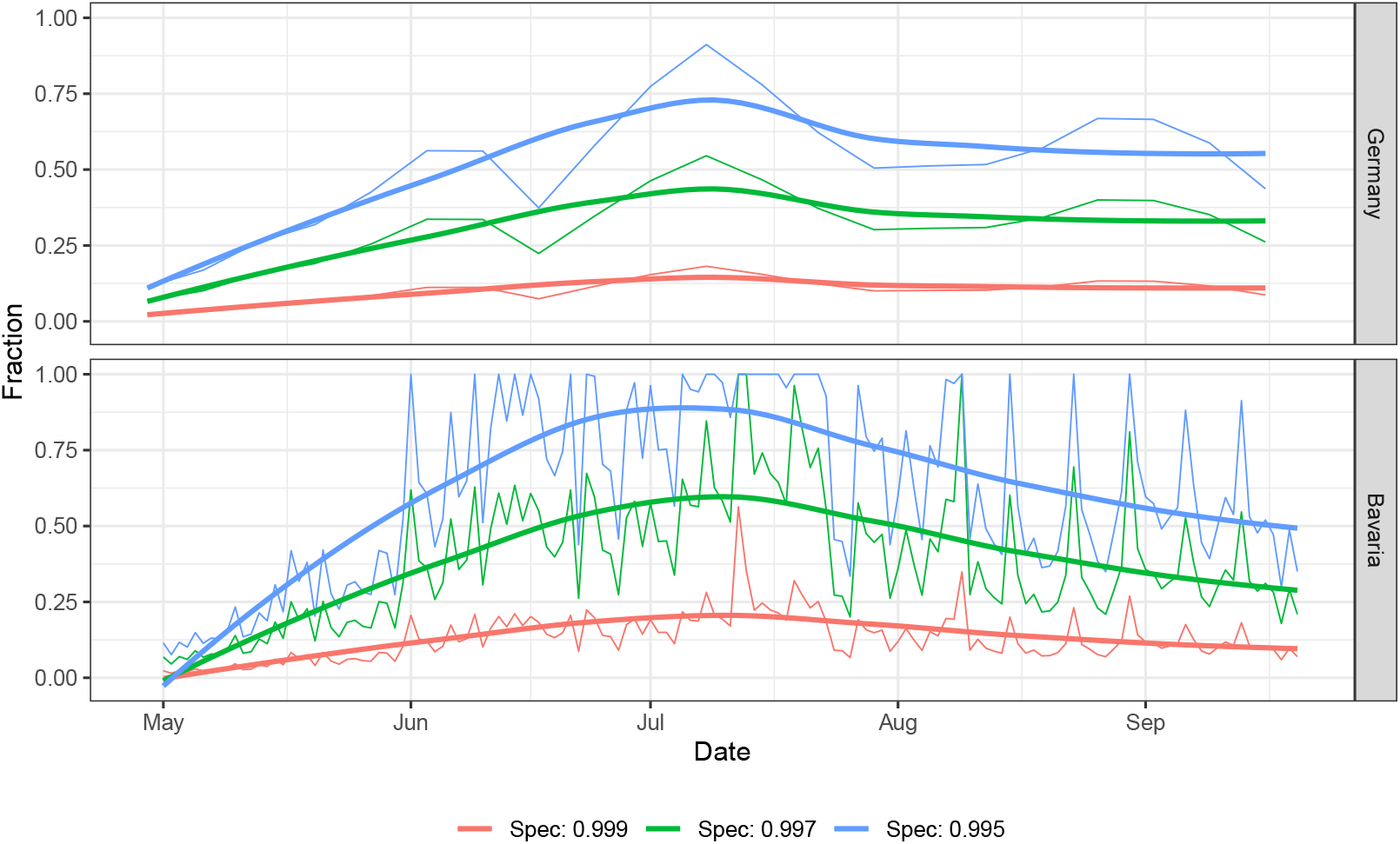
Fraction of false-positives in Germany and Bavaria over time for different assumptions on the specificity. Shown is the fraction of all cases that is removed in the misclassification adjusted case counts due to false-positive cases over time assuming different values for the (time-constant) specificity and a constant sensitivity. For both, German and Bavarian data, the relative effect of the false-positive cases is biggest during July, i.e., in the time of low reported case numbers and rising testing activity and is less important with fewer testing and/or higher case numbers. This fraction is independent of the assumed (constant) sensitivity due to the multiplicative nature of the adjustment for a sensitivity smaller than 100%.

Adjusted case numbers in Germany suggest that, dependíng on the extent of the specificity, the increase in case counts in Beginning of July is indeed partly due to false-positives: adjusted case counts are lower than the observed under the assumption of a sensitivity of 90% However, the increase of cases is still apparent even after the adjustment, which indicates that not the full increase was induced by false-positives. Based on the daily reported Bavarian data, we find a similar pattern. This daily data also prompted us to assume a specificity of no lower than 99.5% Even under the extreme assumption that there were no true cases in June and July and all observed cases were false-positives, the false-positive proportion might not plausibly be larger than 0.5% More precisely, we would have expected more false-positive cases than actually reported in 20 of the 61 days of June and July given the estimated number of examined persons and an assumption of 0.5% false-positive examination results. Of all reported PCR-tests by the Bavarian laboratories in June and July, only 0.56% were reported as positive.

### 3.5 Misclassification-adjusted epidemic curve and time-varying reproduction number in Bavaria

We use misclassification-adjusted case numbers to estimate the epidemic curve or the reproduction number R(t), which provide key information for real-time surveillance. When comparing the epidemic curve adjusted for a diagnostic specificity of 99.7% or 99.5% to the unadjusted epidemic curve, we find structural differences in June/July: while the unadjusted epidemic curve started to rise slowly around Mid-June, the adjusted epidemic curves remained on a very low level until Mid-July (Figure 5A). When additionally accounting for a sensitivity of 90% or 70%, we find the same (Figure 5B); in fact, the impact of different values for the sensitivity was low during this time due to the low incidence. After Mid-July, the increase of the epidemic curve was genuine: in all scenarios, we observe an increase of case counts. In August/September, the reduction of case counts when accounting for false-positives based on a specificity of 99.7% is neutralized by the increase of the case count when considering also false-negatives based on a sensitivity of 70%: the unadjusted and adjusted epidemic curve are basically the same.

**Figure 5:**
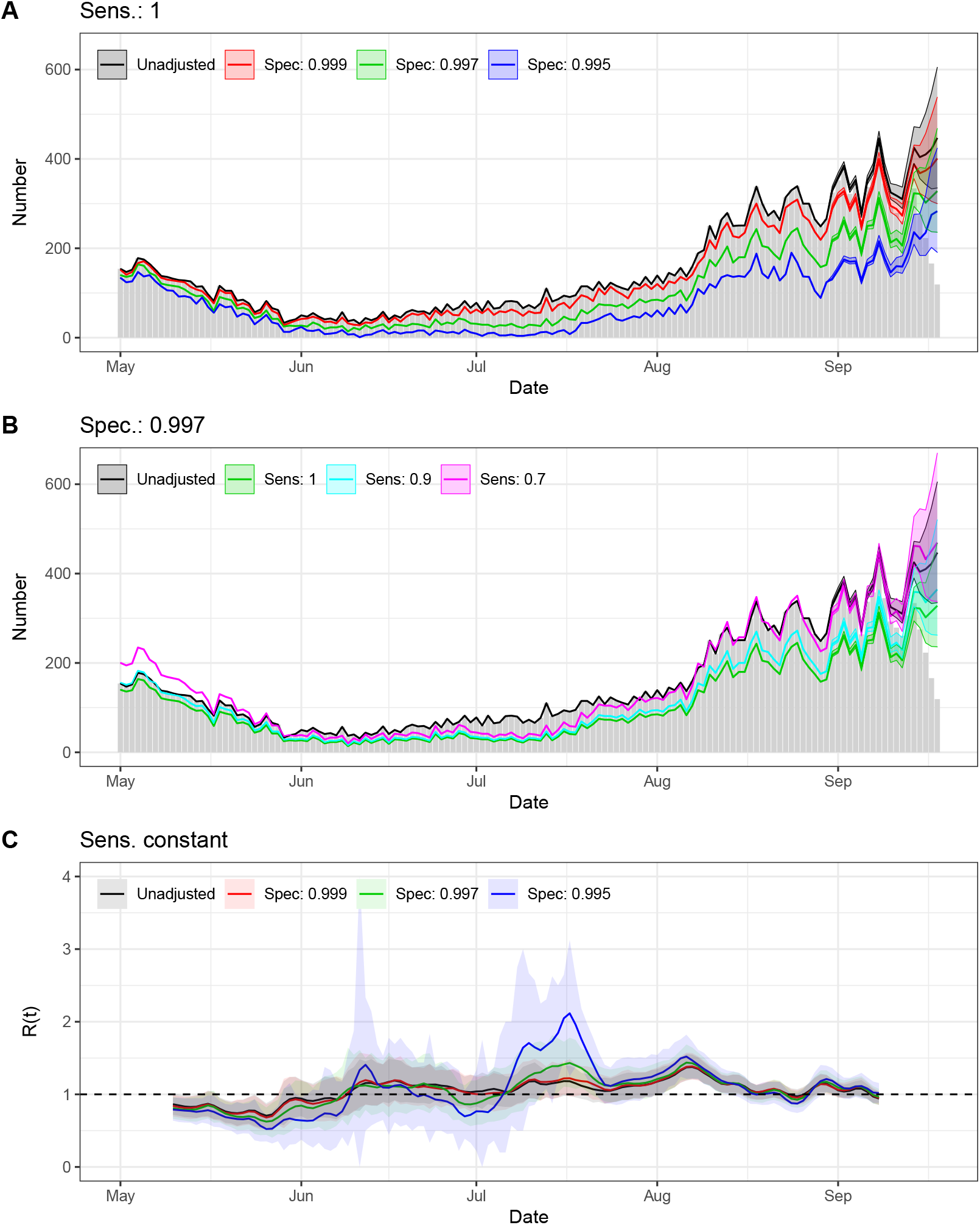
Misclassification adjusted epidemic curve and time-varying reproduction number *R*(*t*). Panel A shows the estimated epidemic curve (number of disease onsets per day) for different assumptions regarding the specificity (assuming a perfect sensitivity as in the unadjusted analysis). Panel B shows the results for different assumptions regarding the sensitivity assuming a specificity of 99.7% and additionally the unadjusted curve. The grey bars show the number of cases with disease onset on a specific day (or imputed if missing) as reported to the LGL until Sept, 21. Panel C shows the misclassification adjusted time-varying reproduction number for different assumptions regarding the specificity. Results are independent of the assumed (time-constant) sensitivity. The epidemic curve is estimated based on a Bayesian hierarchical nowcasting model considering misclassification adjusted case counts, the time-varying reproduction number is estimated based on the estimated epidemic curves.

The misclassification-adjusted estimation of the epidemic curve facilitates estimation of an adjusted time-varying effective reproduction number *R*(*t*). It is an estimate of the average number of individuals that are infected by an individual with disease onset on a given day *t*. If this factor is smaller than one, case numbers are decreasing, if it is bigger than one, case numbers are increasing within the following days. At the beginning of July, adjusted *R*(*t*), is smaller than one when assuming a specificity of 99.5% or 99.7%, while the unadjusted *R*(*t*) was slightly larger than one (Figure 5C). This is in line with the observation that, at that time, true case numbers might have been close to zero or decreasing, but reported numbers were slightly increasing due to false-positives from increased testing. However, shortly thereafter in Mid-July, the adjusted *R*(*t*) exceeds one for all considered values of the specificity, which is in line with increasing case numbers in August/September and an actual true increase in the case counts. The (relative) increase in case numbers in July is bigger when accounting for a relatively high fraction of false-positives in June, yielding bigger estimates of *R*(*t*) the lower the assumed specificity.

## 4 Discussion

In this work, we conceptualize sources of uncertainty in person-specific PCR-based COVID-19 diagnostics and quantify realistic extents of misclassification in terms of plausible values for sensitivity and specificity. We provide an approach to adjust reported case counts for the diagnostic misclassification and extend this approach to a misclassification-adjusted real-time estimation of the epidemic curve and reproduction number R(t). This helps to solve two important problems of case reporting data in real-time surveillance: temporal assignment of reported cases and accounting for misclassification in COVID-19 diagnostics. On the example of data from Germany and Bavaria, we quantify the impact of diagnostic misclassification on the time series of reported case counts and the real-time estimation of the epidemic curve.

Sensitivity and specificity of the diagnostics have a structurally different impact on case counts: a sensitivity smaller than one leads to an underestimation of the true number of cases by a factor independent of the number persons tested, a specificity is smaller than one leads to an overestimation of case numbers, the extent of which depends on the number of tested persons and corresponds to the number of false-positive cases. When the number of tested persons changes over time, the number of false-positives changes as well. This can distort the estimated epidemic curve and thus the apparent dynamics of the epidemic.

For the German case counts during summer 2020, we find that the reported case numbers during the low incidence phase in June and July were to a relevant part false-positives, but that the observed increase of cases since Mid-July was not entirely driven by false-positives. When accounting for false-positive and false-negative test results, the adjusted case numbers in August and September were on a similar level than the unadjusted. Therefore, the increase seen in German case numbers from July until September, which was debated as being predominantly driven by false-positives due to increased testing activity, was a genuine increase of infections. Based on our developed analysis approach such questions can also be answered in future real-time surveillance.

Since the relative impact of misclassification on reported case counts depends on the number of examined persons and the current incidence, we update our analyses on a regular basis and provide the results adjusted for various scenarios of sensitivity and specificity on our website (https://corona.stat.uni-muenchen.de/nowcast/). Based on current results from January, 2021, it becomes obvious that the true number of cases during the strong second wave in Bavaria might be considerably underestimated due to false-negative results (Supplemental Figure 2). The general structure of the epidemic curve is, however, not changed by taking into account errors in COVID-19 examinations. The distortion due to false-positive tests hardly plays a role in times of high incidence.

Our analyses have some assumptions and limitations. The adjustment for misclassification in COVID-19 diagnostics depends on accurate information with respect to the number of examined individuals. Such information is not directly available in German surveillance data and we rely on a model-based approach to estimate this number. The results appear plausible, but cannot be directly validated. Furthermore, we assume constant misclassification probabilities for the COVID-19 diagnostics over time. This assumption is likely violated by changes in the diagnostic procedures, changes in workload for the laboratories, and changes or improvements in standard operating procedures. We believe, however, that our calculations over a range of plausible assumptions for sensitivity and specificity can give a realistic overview about potential biases due to misclassification.

Misclassification in COVID-19 diagnostics is not the only problem for the interpretation of reported case numbers. It is well known that not all infected persons are captured and examined. This leads to a relevant difference between the number of infected individuals and the number of reported infections. This problem is not COVID-19-specific, but also occurs with other diseases when they only cause mild symptoms in some cases - as typically described by the so-called surveillance pyramid [24]. The case detection ratio for COVID-19 was estimated in Germany from antibody prevalence studies to be as low as 0.2 to 0.4 for the first phase of the pandemic in spring 2020 [25, 26, 27]. If the case detection ratio is constant over time, the structure of the epidemic curve remains unchanged. However, expanding the testing activity increases the detection ratio. Therefore, one has to be careful when comparing absolute numbers of cases over longer periods of time. More specifically, the absolute number of reported cases in Germany during March-April 2020 is not comparable to the case numbers reported during August and September (and today) due to increased testing. In our analysis and figures, we focused on the epidemic curve starting in May 2020, i.e., after the first phase of the pandemic. Nevertheless, the remaining increase of the misclassification-adjusted case numbers during August-September might still partly be driven by an increasing case detection ratio. This question cannot be answered directly from reported case numbers, but requires additional information, e.g. numbers of hospital admissions, deaths or longitudinal data on antibody prevalence. Recent modelling approaches [28] might also be extended to account for misclassification in COVID-19 testing.

Overall, a thorough analysis of case reporting data adjusting for misclassification is important to improve monitoring of short-term changes in the pandemic situation. Given the high relevance of reported case counts for the surveillance of an acute pandemic, we recommend to analyse case counts adjusted for plausible assumptions regarding diagnostic misclassification in the corresponding testing regimes to understand potential distortions due to misclassification and avoid the appearance of false precision. This remains especially important as containment and surveillance of the current pandemic remains a central task worldwide. Consideration of misclassification in tests will also be relevant with the increasing use of alternative test methods, especially if their results are not confirmed by the established PCR-tests. In this work, we have shown that a thorough analysis of surveillance data can help to capture current trends more reliably and reduce the ambiguity of the available information, which can ultimately support public confidence in the available evidence. This will also be important in the event of a future emergence of new virus strains or other pandemic pathogens.

## Data Availability

Code for reproducing our analysis and adapting it to other application scenarios is available at Github. There, we also provide an artificial data set on the person-specific case reporting data from Bavaria.

https://github.com/FelixGuenther/mc_covid_cases_public

