## Supplemental Material for "Analysis of COVID-19 case numbers: adjustment for diagnostic misclassification on the example of German case reporting data"

### Supplemental Note 1 - Misclassification adjustment for case numbers

We denote a binary indicator variable $D_i$ for the event that a person $i$ is currently infected. $D_i = 1$ means that a person is currently infected by COVID-19 and $D_i = 0$ that a person is not infected. The result of the person's COVID-19 examination (e.g., one or multiple sequential PCR-tests) is denoted by $T_i$. $T_i = 1$ corresponds to a classification as infected (positive examination) and $T_i = 0$ to a negative COVID-19 examination. Furthermore, the sensitivity (i.e., the probability of a true positive examination) is defined by sens $= P(T_i = 1|D_i = 1)$ and the specificity (probability of true negative examination) is denoted by spec $= P(T_i = 0|D_i = 0)$. Then the probability of a positive examination is given by

$$P(T_i = 1) = P(D_i = 1) \cdot \text{sens} + P(D_i = 0) \cdot (1 - \text{spec}). \tag{1}$$

We denote the (unknown) number of examined persons at a certain time $t$ by $NT_t$ and the number of positive diagnostic procedures (i.e., reported cases) by $T_t^+ = \sum_{i=1}^{NT_t} T_i$. We can rewrite the expected number of reported cases on day $t$ as

$$E(T_t^+|D_t^+, NT_t) = D_t^+ \cdot \text{sens} + (NT_t - D_t^+) \cdot (1 - \text{spec}), \tag{2}$$

where $D_t^+$ denotes the (unknown) number of actually infected persons that are examined on day $t$. Equation (2) is derived from equation (1) by replacing the probabilities $P(\cdot)$ by the corresponding relative frequencies, $P(T_i = 1) = T_t^+/NT_t$, $P(D_i = 1) = D_t^+/NT_t$, and $P(D_i = 0) = (NT_t - D_t^+)/NT_t = D_t^-/NT_t$, and multiplying both sides with the number of examined persons, $NT_t$.

Equation (2) shows that the effects of sensitivity and specificity on the observed case counts are different. While a low sensitivity leads to an underestimation of the number of cases by the factor sens, the effect of specificity is additive and depends on the number of examined persons.

If all information, i.e., the number of reported cases $T_t^+$, the number of examined persons $NT_t$, and the sensitivity and specificity of the person-specific examination are known, we can rewrite equation (2) and estimate the number of true cases $D_t^+$ based on

$$\hat{D}_t^+ = \frac{T_t^+ - NT_t \cdot (1 - \text{spec})}{\text{sens} + \text{spec} - 1}. \tag{3}$$

*

This estimator relates to the well known matrix method, see e.g., Rogan and Gladen (1978).

Since only positive COVID-19 examinations are directly reported to German health authorities, only the overall number of reported cases $T_t^+$ is directly available, but the corresponding number of examined individuals, that would be reported as case on day $t$ in the event of a positive test, $NT_t$, is unknown. However, the overall number of performed tests is separately reported by the laboratories as well as the number of positive tests. This data can have a difference in temporal allocation and, furthermore, we expect more reported tests than examined persons due to multiple tests for single persons and - in case of the Bavarian data - tests of individuals living outside of Bavaria.

To establish the relationship between the two quantities, we utilize the positive test results and model the number of reported cases at the health authorities based on the number of positive tests reported by the laboratories from the current and previous time points (i.e., based on lagged time series of reported positive tests from the laboratories). We utilize two different models and consider different degrees for the lag-number of positive tests. The first model corresponds to standard linear regression with linear effects of the (lagged) number of positive tests from the same and previous time points:

$$E(T_t^+) = \sum_k \alpha_k \cdot \text{Test}_{t-k}^+, \quad t = \text{May 1st}, \ \ldots, \ T. \tag{4}$$

Here, the number of positive tests reported by the laboratories at time point $t$ is denoted by $\text{Test}_t^+$ and T corresponds to the most current time point. We estimate the model based on all possible subsets of lags $k \subset \{0, \ldots, 7\}$. As a second model, we consider a varying-coefficient model (Hastie and Tibshirani, 1993), in which the linear effect $f_k$ of the (lagged) number of positive tests, $PCR_{t-k}^+$, varies smoothly over time:

$$E(T_t^+) = \sum_k f_k(t) \cdot \text{Test}_{t-l}^+, \quad t = \text{May 1st}, \ \ldots, \ T. \tag{5}$$

This model is estimated for all different subsets of lags $k \subset \{0, \ldots, 7\}$, as well. From all estimated models, we select the best performing model based on the Bayesian information criterion (BIC).

Assuming a similar relation between the number of (positively) reported tests for individuals reported as cases to the Bavarian/German health authorities as for the number of reported tests per examined persons, we estimate the number of examined persons $NT_t$ by using the estimated parameters of the selected model (4) or (5) and plug-in the total number of reported tests:

$$\widehat{NT}_t = \sum_k \hat{f}_k(t) \cdot \text{Test}_{t-k}, \tag{6}$$

where $\text{Test}_t$ denotes the total number of test reported by the laboratories on day $t$ and $\hat{f}_k(t)$ are the estimated associations for the (lagged) number of tests reported by the laboratories. Depending on whether a model of type (4) or (5) is selected, they correspond to time-constant (linear) effects $f_k(t) = \alpha_k$ or (linear) effects that vary smoothly over time.

Plugging the observed numbers of positively examined persons (new cases), $T_t^+$, and the derived number of relevant examinations, $\widehat{NT}_t$ from (6), into (3), we obtain misclassification adjusted estimates for the number of new COVID-19 cases per time point, $\hat{D}_t^+$.

In the nowcasting, we estimate the number of cases with disease onset on a specific day based on a complex Bayesian hierarchical model using individual-specific data on the reporting and disease onset date (cf. Günther et al. (2020)). To apply the proposed adjustment for misclassification, we first focus on the scenario of no false-negative examinations (sensitivity equals one) and a reduced specificity smaller than one. We then calculate the expected number of false positives reported to the health authorities on a certain day based on the difference of

$$\hat{D}_t^+ = \frac{T_t^+ - NT_t \cdot (1 - \text{spec})}{\text{spec}} \tag{7}$$

and the reported number of new cases $T_t^+$. Then, we remove this number of randomly selected observations from our data and apply the nowcasting to the reduced data to estimate the *false-positive adjusted* epidemic curve. Based on the estimated epidemic curve it is possible to estimate the effective time-varying reproduction number $R_e(t)$ as also described in Günther et al. (2020).

To take possible false negatives into account, we can rewrite formula (3) by

$$\hat{D}_t^+ = \frac{T_t^+ - NT_t \cdot (1 - \text{spec})}{\text{spec}} \cdot \frac{\text{spec}}{\text{sens} + \text{spec} - 1} \tag{8}$$

and plug in different values for the sensitivity, $\text{sens} < 1$. The first term of (8) corresponds to the false positive adjusted estimate from (7) and the second part is a constant factor independent of the number of examinations per day.

Since the *false-negative* adjustment relies on a constant factor which is independent of the number of tested individuals, the factor $\text{spec}/(\text{sens} + \text{spec} - 1)$ can be directly applied to the result of the false positive adjusted nowcasting procedure, which reduces the computational effort considerably compared to a repeated application of the nowcasting to (upsampled) data. Note, that $\text{spec}/(\text{sens} + \text{spec} - 1) \approx 1/\text{sens}$ for a specificity close to one. The false negative adjustment corresponds therefore roughly to a point-wise up-scaling of the estimated epidemic curve by the reciprocal sensitivity.

### Supplemental Note 2 - Results of model for approximating the number of examined persons over time

To approximate the number of examined persons per time interval (week or day) based on the reported number of performed PCR-tests by the laboratories, we estimate a model that establishes a relation between the number of reported cases by the health authorities and the number of reported positive PCR-tests by the laboratories also allowing for lagged associations. We select the best-performing model out of a set of candidate models with time-varying or time-constant linear associations between the number of reported cases and the (lagged) number of positive PCR-tests based on the Bayesian information criterion (BIC) (cf. Supplemental Note 1). With this approach, we aim at capturing potentially time-varying associations between the available information in a flexible way to obtain a plausible approximation for the number of examined individuals over time. Nevertheless, a closer look at the estimated associations might help to understand the structure in the available data better.

For the weekly German data, a varying coefficient model was selected with time-varying effects of the number of reported PCR-tests in the same week and the previous week. For the daily Bavarian data, the selected model considers time-varying effects of the number of positive PCR-tests on the respective day and additionally of the number of positive PCR-tests two and six days before.

At a given time $t$, the predicted number of reported cases at the health authorities corresponds to a weighted sum of reported positive PCR-tests at the same time and previous time points (i.e., previous week for the German data, and day $t-2$ and $t-6$ for the Bavarian data). The weights are given by the estimated effect for time point $t$. Analogously, our predicted number of examined persons corresponds to the weighted sum of reported PCR-test counts (positive and negative).

For the German model, we find a positive association between the number of reported cases by the health authorities and the number of reported positive PCR-tests in the same week that increases over time towards one. Additionally, we estimated a small positive contribution (around 10%) of the reported positive PCR-tests in the previous week. The rather low weight of the lag-0 PCR-tests in the beginning of the observation period corresponds to the observation that the German laboratories reported more positive PC-tests to the health authorities during May and June than new cases were reported by the RKI (cf. Figure 1 in the manuscript). This indicates that several laboratories reported tests that were already performed at in previous weeks (i.e., during the first wave) in this time period. During the end of the observation period PCR-test reporting by the laboratories was better established and the weekly number of reported positive tests corresponds closely to the number of reported cases by the RKI. In fact, for the German data the misclassification adjustment in August/September is very similar when utilizing the number of reported tests instead of the predicted number of examined persons (not shown).

For the Bavarian model estimated based on daily data, we obtain somewhat similar results. The association of the number of reported positive PCR-tests and the number of reported cases at the health authorities per day increases over time towards around one. Additionally we have a small contribution of the number of positive PCR-tests two and six days before the current day that remains rather constant over time. The lag-0 association (i.e., the association of the number of reported positive tests and the number of reported cases at the same day) shows, however, an additional change in July and August. At this time, there were more positive tests reported by the Bavarian laboratories than cases reported by the Bavarian health authorities and the lag-0 association decreases slightly. This pattern could be related to the German holidays, when many travellers from other federal states were tested when passing through Bavaria. At the end of the observation period the effect of the number of positive tests reported by the laboratories is slightly bigger than one: there are (already) more cases reported by the health authorities than positive tests reported by the laboratories. This might be induced by a faster reporting of person-specific positive test results to the local health authorities (leading to a case registration) compared to the reporting of aggregated (positive and overall) test-numbers by the laboratories. This indicates that the approximation of the number of examined persons based on the flexible model is an important step for a valid misclassification adjustment, especially when analysing daily data in close to real-time.

### Supplemental Figures

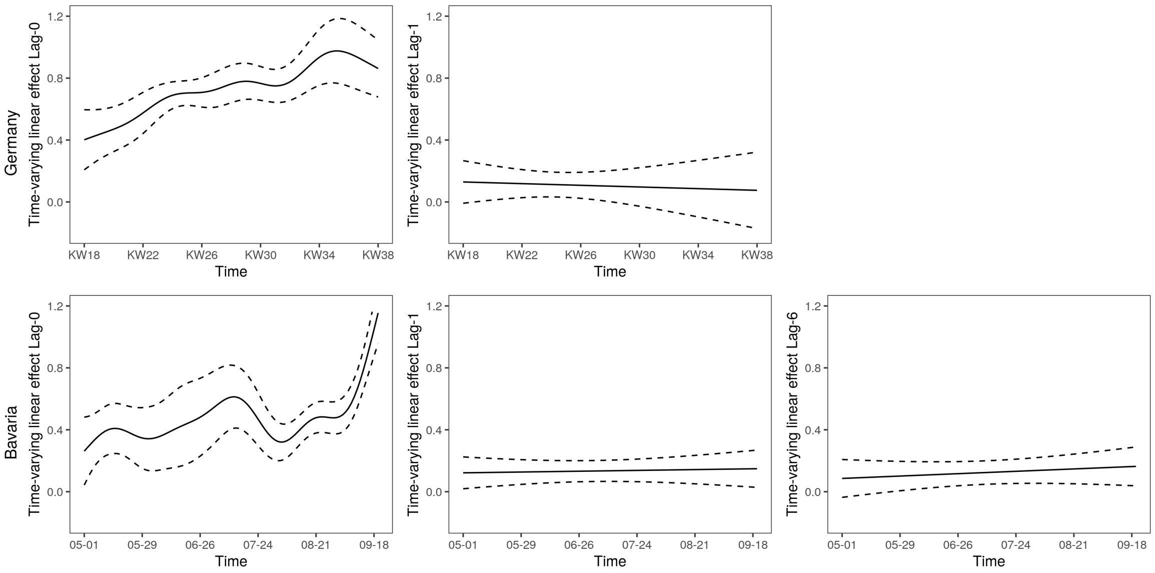


Supplemental Figure 1: Estimated time-varying association between (lagged) numbers of positive tests reported by the laboratories and the number of reported cases at the health authorities. The model for Germany (top row) is based on weekly information and contains time-varying effects of the number of reported positive tests in the current and the previous week, the model for Bavaria (bottom row)is based on daily information and contains time-varying effects of the number of reported positive tests at the current day, and two and six days before.

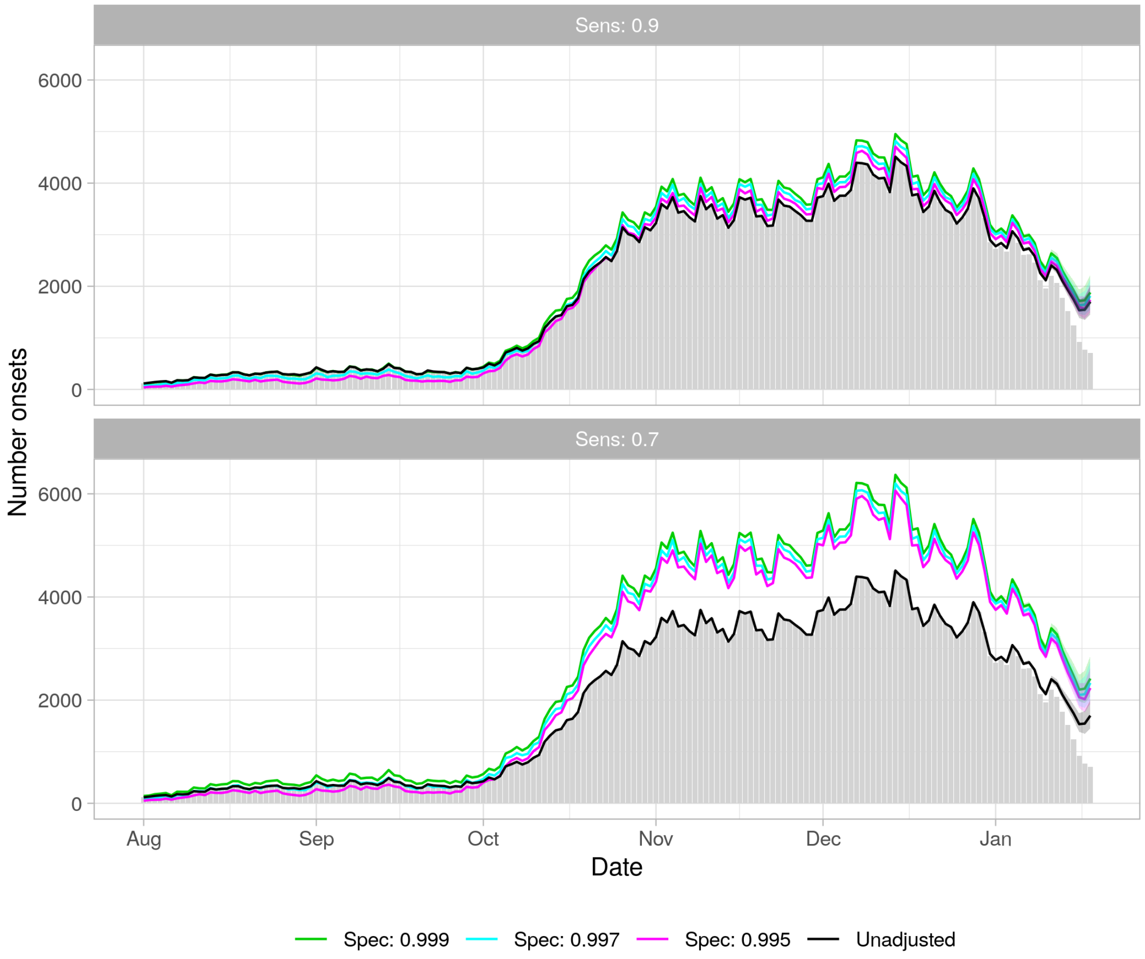


Supplemental Figure 2: Estimated misclassification-adjusted epidemic curve for Bavaria based on data available an January, 21, 2020. They grey bars show the number of disease onsets per day reported until January, 21. The black line represents the estimated epidemic curve from nowcasting without adjustment for misclassification. The colored lines show the estimated epidemic curve misclassification adjustment based on different assumptions regarding the sensitivity and specificity of person-specific COVID-19 examination. While the true number of cases might be considerably underestimated due to false-negative results (Supplemental Figure 2), the general structure of the epidemic curve is not changed by taking misclassification into account.
